# Using Meta-Transformers for Multimodal Clinical Decision Support and Evidence-Based Medicine

**DOI:** 10.1101/2024.08.14.24312001

**Authors:** Sabah Mohammed, Jinan Fiaidhi, Abel Serracin Martinez

## Abstract

The advancements in computer vision and natural language processing are keys to thriving modern healthcare systems and its applications. Nonetheless, they have been researched and used as separate technical entities without integrating their predictive knowledge discovery when they are combined. Such integration will benefit every clinical/medical problem as they are inherently multimodal - they involve several distinct forms of data, such as images and text. However, the recent advancements in machine learning have brought these fields closer using the notion of meta-transformers. At the core of this synergy is building models that can process and relate information from multiple modalities where the raw input data from various modalities are mapped into a shared token space, allowing an encoder to extract high-level semantic features of the input data. Nerveless, the task of automatically identifying arguments in a clinical/medical text and finding their multimodal relationships remains challenging as it does not rely only on relevancy measures (e.g. how close that text to other modalities like an image) but also on the evidence supporting that relevancy. Relevancy based on evidence is a normal practice in medicine as every practice is an evidence-based. In this article we are experimenting with meta-transformers that can benefit evidence based predictions. In this article, we are experimenting with variety of fine tuned medical meta-transformers like PubmedCLIP, CLIPMD, BiomedCLIP-PubMedBERT and BioCLIP to see which one provide evidence-based relevant multimodal information. Our experimentation uses the TTi-Eval open-source platform to accommodate multimodal data embeddings. This platform simplifies the integration and evaluation of different meta-transformers models but also to variety of datasets for testing and fine tuning. Additionally, we are conducting experiments to test how relevant any multimodal prediction to the published medical literature especially those that are published by PubMed. Our experimentations revealed that the BiomedCLIP-PubMedBERT model provide more reliable evidence-based relevance compared to other models based on randomized samples from the ROCO V2 dataset or other multimodal datasets like MedCat. In this next stage of this research we are extending the use of the winning evidence-based multimodal learning model by adding components that enable medical practitioner to use this model to predict answers to clinical questions based on sound medical questioning protocol like PICO and based on standardized medical terminologies like UMLS.

## I. Introduction

Meta-Transformers have demonstrated significant promise for computer multimodal learning tasks involving variety of data from natural language, 2D images, 3D point clouds, audio, video, time series and tabular data [1]. Multimodality learning is the new AI challenge to develop models that have the ability to learn simultaneously from different sources of information [2]. In recent years models such as Next-GPT [3], CLIP [4], Flamingo [5], VLMO [6], OFA [7] and BEiT-3 [8] reported promising progress in processing text and image inputs compared to single-modality model. However, there are a lot of work still remains to be done to extend multimodal learning models to different domains as well as to enhance its accuracy. There are many reasons hindering the progress towards having unified learning network for processing various modalities. Such reasons include the difficulty of integrating of the attention mechanisms [9], the difficulty to identify good fine-tuning mechanisms [10] and lack of providing evidence-based predictions [11]. Most of the current research investigations to overcome these difficulties are focused on identifying generic techniques that can associate text and images via variety of embedding techniques as text and images are the most common forms to describe clinical cases. These generic learning techniques are trained using a contrastive learning approach that aims to unify text and images, allowing tasks like image classification to be done with text-image similarity. The core architecture of all these generic techniques consist of a text encoder and an Image encoder where they are jointly trained to predict the correct pairings of a batch of training (image, text) samples without being trained for a specific domain and may requires little fine-tuning to improve its accuracy. However, generic learning models like CLIP trained using 400 million (text and image) pairs from the ImageNet has not shown promising results in the medical domain [12] and requiring intensive fine tuning to reach SOTA performance. Attempts to redefining the CLIP learning model for medical use has been divided into three groups: (1) Medical Knowledge Enhancement Approach for Subject and Domain Levels [13], (2) Supervision Focused Approach within particular modality and across modalities [14] and (3) Semantic-based Pre-Training Approach [15]. All these approaches aim to improve training the text-image training for medical domain but it does not necessary build the association and similarity utilizing evidence-based medical sources like PubMed [16]. In this article we are investigating meta-transformer generic learning technologies that can be used to support evidence-based medicine (EBM) where it can be used reliably in diagnosis and prognosis of described clinical cases.

## II. Medical Meta-Transformers for EBM

Meta-transformers are special type of learning transformers tuned to work with multimodal medical data [17]. In this direction, the meta-transformers need to work with multimodal datasets like those listed in table 1.

**Table 1:**
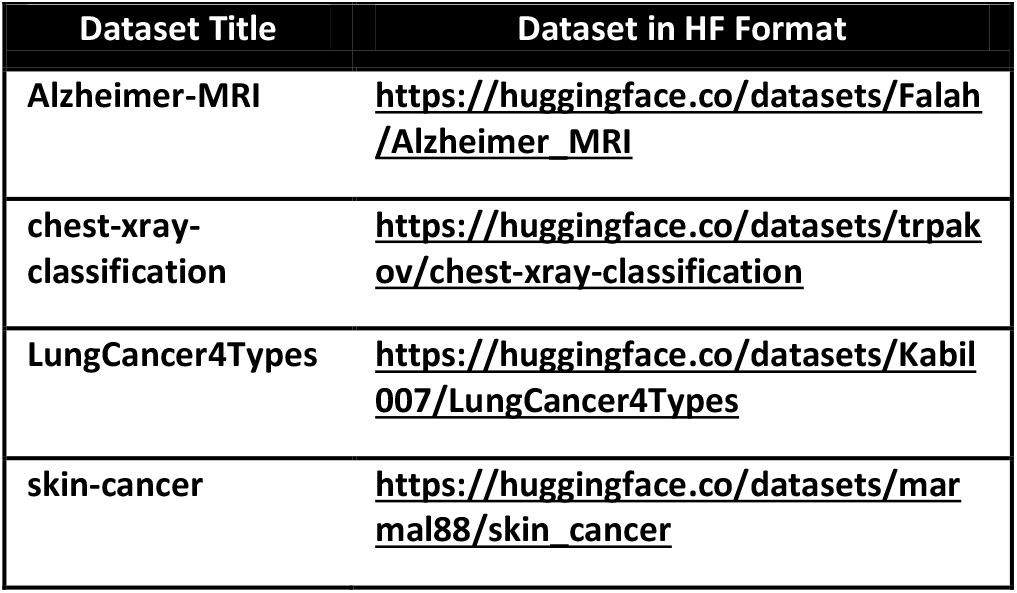
Common Medical Datasets.

However, we can extend the list of datasets by uploading other datasets from the Hugging Face hub^2^ or by uploading your own dataset to the Hugging Face hub then import it: https://huggingface.co/docs/datasets/en/share#create-the-repository

Actually the Hugging Face Hub hosts many machine learning models and datasets for a broad range of tasks across textual narratives (NPL), imaging (Computer Vision), and Audio. Not all models are pre-trained for medical domains nor they are built from transformers. For this purpose, there is a need to select models that are pretrained on medical domains as well as follows similar encoding standard like the Open AI CLIP^3^ where these

models have the ability to connect textual narratives via a text encoder and pair it with related images based on image encoder. This type of text-image pairing is not only important for multimodal learning but also important to answer clinical questions [18]. The following Python snippet illustrates using Open AI CLIP and the BRACS^4^ dataset [19] to list the different histological disorders.

import numpy as np

import torch

import open_clip

open_clip.list_pretrained()

import os

import histolab.data

import IPython.display

import matplotlib.pyplot as plt

from PIL import Image

import numpy as np

from collections import OrderedDict import torch

%matplotlib inline

%config InlineBackend.figure_format = ‘retina’

descriptions = {

“N”: “Normal Tissue”,

“PB”: “Pathological Benign”, “UDH”: “Usual Ductal Hyperplasia”, “FEA”: “Flat Epithelial Atypia”,

“ADH”: “A Typical Ductal Hyperplasia”, “DCIS”: “Ductual Carcinoma in Situ”, “IC”: “Invasive Carcinoma”,

“UN”: “Unknown”

}

original_images = []

images = []

texts = []

plt.figure(figsize=(16, 5))

for filename in [filename for filename in

os.listdir(bracs.data_dir) if filename.endswith(“.png”) or

filename.endswith(“.jpg”)]:

name = os.path.splitext(filename)[0]

if name not in descriptions:

continue

image = Image.open(os.path.join(bracs.data_dir,

filename)).convert(“RGB”)

plt.subplot(2, 4, len(images) + 1)

plt.imshow(image)

plt.title(f”{filename}\n{descriptions[name]}”)

plt.xticks([])

plt.yticks([])

original_images.append(image)

images.append(preprocess(image))

texts.append(descriptions[name])

plt.tight_layout()

The result from this simple code snippet illustrates associating tissue disorders with their textual label (see Table 2).

**Table 2:**
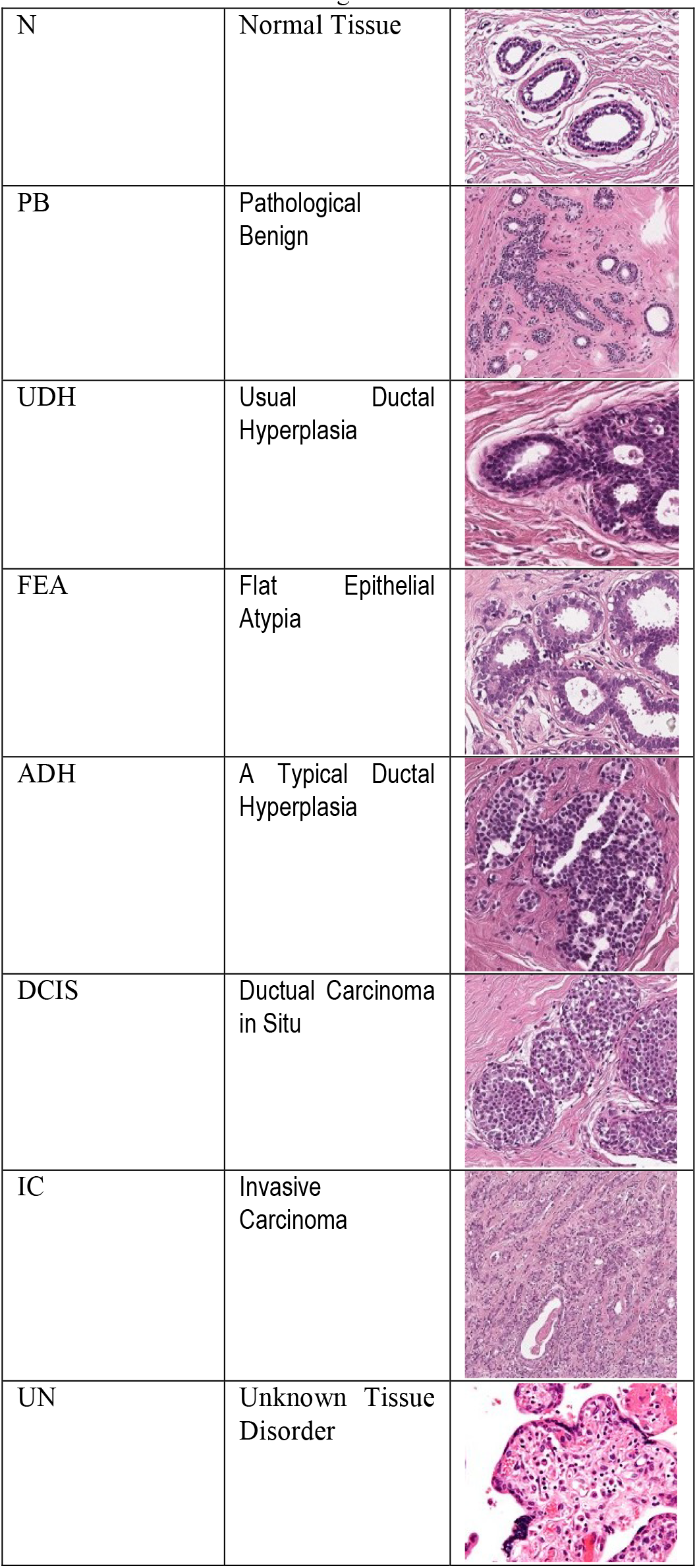
Different Histological Tissue Conditions.

|  |  |
| --- | --- |
| N | Normal Tissue |
| PB | Pathological Benign |
| UDH | Usual Ductal Hyperplasia |
| FEA | Flat Epithelial Atypia |
| ADH | Atypical Ductal Hyperplasia |
| DCIS | Ductal Carcinoma in Situ |
| IC | Invasive Carcinoma |
| UN | Unknown Tissue Disorder |

However, training learning models on any given medical dataset like BRACS requires that these models which are originally pre-trained on general medical domain data need to be fine-tuned on the specific medical given domain like the skin-cancer. In this direction, the Hugging Face hub contains models that are general trained on medical domains like:

### Pubmed CLIP

https://huggingface.co/flaviagiammarino/pubmed-clip-vit-base-patch32

### ClipMD

https://huggingface.co/Idan0405/ClipMD

### BiomedCLIP-PubMedBERT

https://huggingface.co/microsoft/BiomedCLIP-PubMedBERT_256-vit_base_patch16_224

### BioCLIP

https://huggingface.co/imageomics/bioclip

These pre-trained models can perform very well on specific tasks and datasets, but they do not generalize well. They cannot handle new classes or images beyond the domain they have been trained with [20]. Retraining and fine-tuning is an option, as training requires significant time and capital investment for gathering a classification dataset and the act of model training itself [21]. There are a few reasons:

### Saves resources

Training a large model from scratch requires significant computational resources and time. By using a pre-trained model, we can leverage the patterns it has already learned, reducing the resources required.

### Leverages transfer learning

This is a big part of fine-tuning. The idea is that the knowledge gained while solving one problem can be applied to a different but related problem.

### Deals with limited data

In many cases, we might not have a large enough dataset for our specific task. Fine-tuning a pre-trained model on a smaller dataset can help prevent overfitting, as the model has already learned general features from the larger dataset it was initially trained on.

Fortunately, OpenAI’s CLIP models have proved itself as incredibly flexible learning models that often require zero retraining. Models like Pubmed CLIP and BioCLIP are called “*many-shot*” learning models because we need many training samples to reach acceptable performance during that final fine-tuning step. Many-shot learning is only possible when we have compute, time, and data to allow us to fine-tune our models. Ideally, we want to maximize model performance while minimizing N-shot requirements. *Zero-shot* is the natural best-case scenario for a model as it means we require zero training samples before shifting it to a new domain or task [22]. Moreover, OpenAI’s CLIP models are initially trained on a dataset called WebImageText (WIT)^5^. WIT contains 400 million (image, text) pairs from publicly available internet data including those covering medical domains. However, OpenAI CLIP models may not be breaking SotA performance benchmarks on specific datasets but still, it is proving to be a massive leap forward in zero-shot performance across various tasks in both image and text modalities [23].

To facilitate experimentation with these medical OpenAI CLIP models, we will need a platform that enables the flexible models changing as well as the flexibility to add or delete datasets. In this direction we found the TTi-Eval Embedding API providing a flexible platform to test the performance of variety of OpeAI CLIP models and datasets:

https://github.com/encord-team/text-to-image-eval?tab=readme-ov-file

All what we need to install this platform is two libraries:

!pip install poetry

!poetry run tti-eval list –all

Algorithm 1 illustrates how we can use the TTi-Eval platform with the four types of transfer learning across different medical datasets.

#### Algorithm 1

Train and Evaluate Models and Datasets

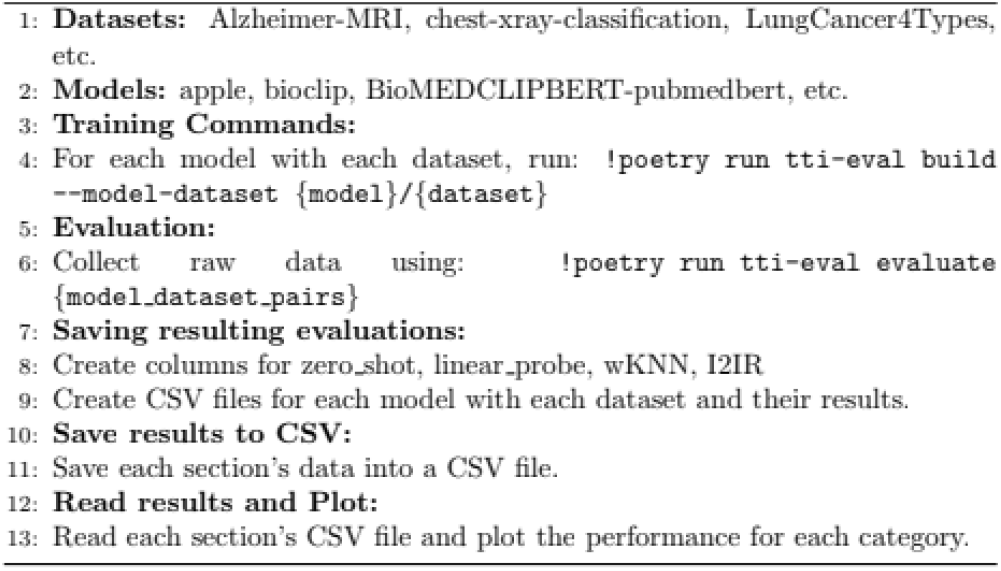

Actually the TTi-Eval platform enable us not only to accommodate different OpenAI CLIP medical models and medical datasets, it also provide us with four basic transfer learning mechanisms that can be used across different datasets including Zero-Shot, Linear-Probe, wKNN and I2IR. Basic to all these transfer learning models are the image encoders and the text encoder. The first image encoder uses the ResNet-50 as base model. The second image encoder considered is Vision Transformer, ViT. The text encoder is a Transformer. Figure 1 describe the performance results of running four OpenAI CLIP models (BioCLIP, CLIPMed, PubmedCLIP and BioCLIP-PubmedCLIP) across four medical datasets (Alzheimer-MRI, chest-xray-classification, LungCancer4Types and skin-cancer).

**Fig. 1:**
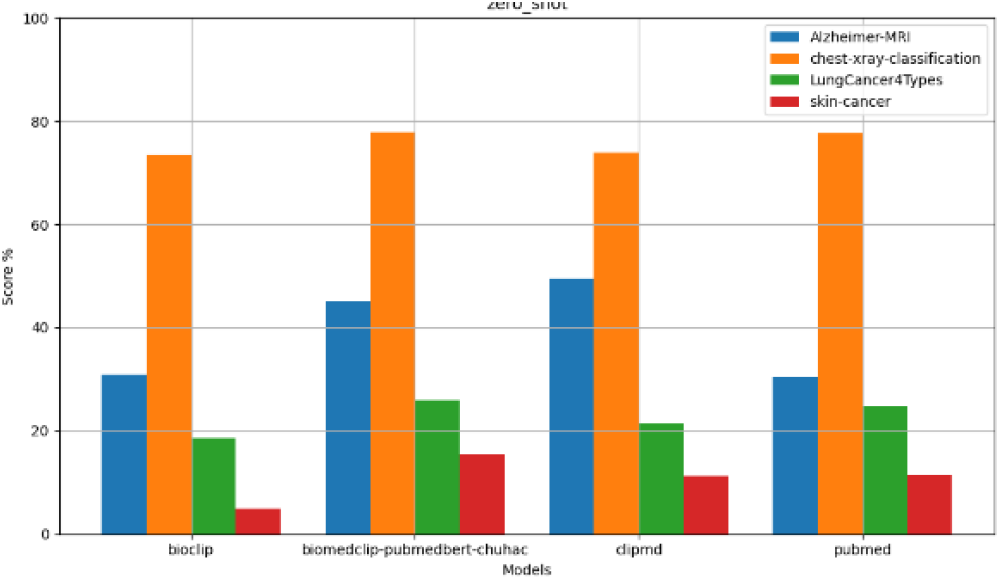
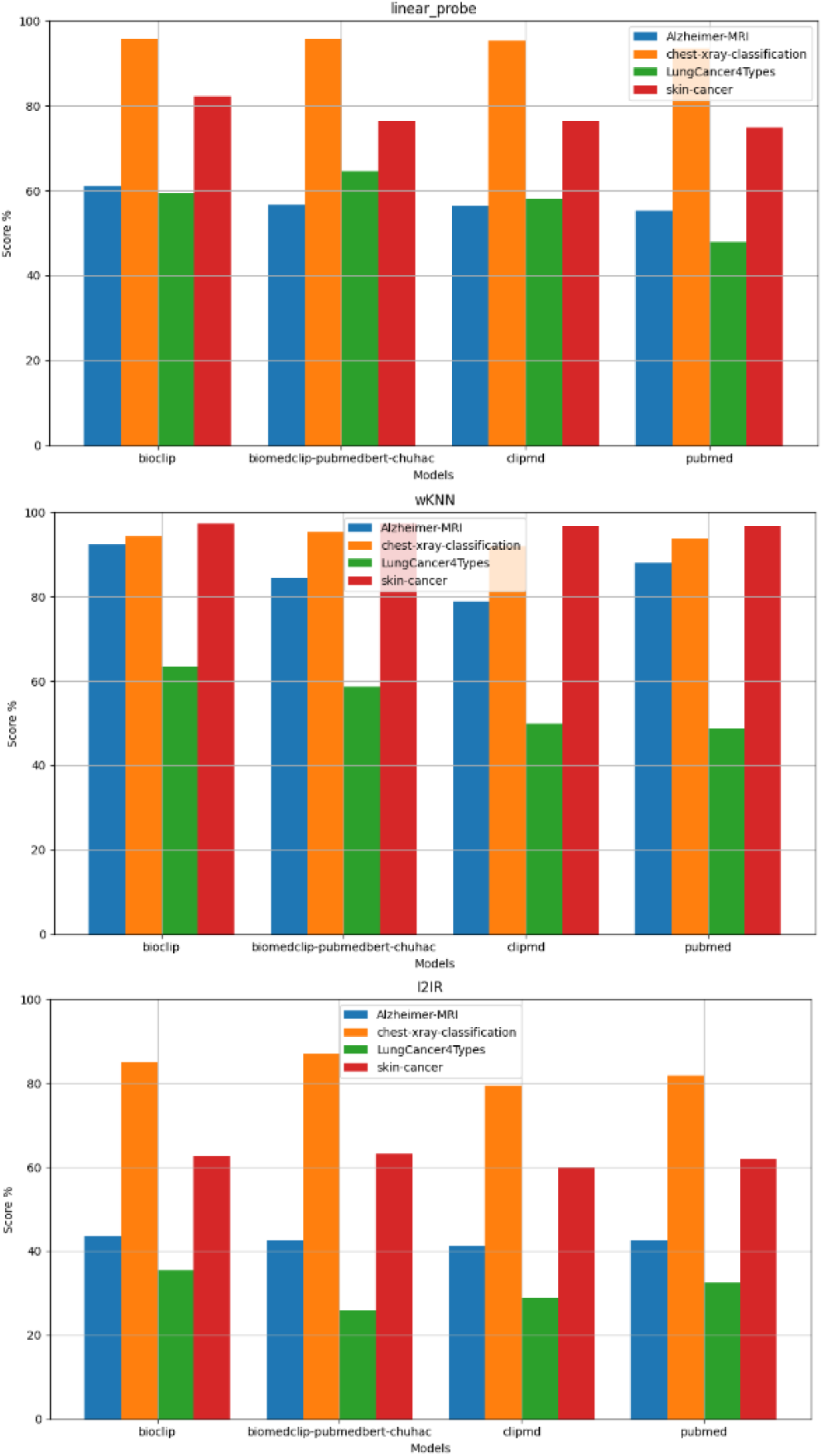
Testing the Performance of Four OpenAI CLIP Models across Four Datasets.

From figure 1, we can see that the overall best performing models is the BiomedCLIP-PubMedBERT over mainly two datasets; the chest-xray-classification and skin-cancer dataset. Further justifications for the wining BiomedCLIP-PubMedBERT model are listed below:

### 1. Balanced Across Multimodal Tasks

While some of the other models may excel in specific medical tasks but falter in others, BiomedCLIP-PubMedBERT maintains a consistently high level of performance across multiple modalities.

### 2. Zero Shot Capabilities

Even in zero-shot scenarios, where the model has not been fine-tuned on the specific medical domain downstream tasks, BiomedCLIP-PubMedBERT performs very well.

### 3. Provides Evidence Based Authenticity

BiomedCLIP-PubMedBERT uses multimodal data from high-quality clinical research that is centerlly publish in the PubMed platform. PubMed is very popular Evidence-Based publication platform and hence BiomedCLIP-PubMedBERT model provides better likelihood of good evidence-based embeddings compared to the other models.

However, we attempted to test the BiomedCLIP-PubMedBERT model further by performing another randomized test using ten cases (images + captions) from the ROCO V2 dataset [24]. Figure 2 illustrates the cosine similarity of testing BiomedCLIP-PubMedBERT on ten cases listed in table 2. Figure 2 prove that the BiomedCLIP-PubMedBERT can reliably identify which caption matches the correct image with higher reliability than other cases. Algorithm 2 illustrates how are testing the different meta-transformers using randomized samples from ROCO V2.

**Fig. 2:**
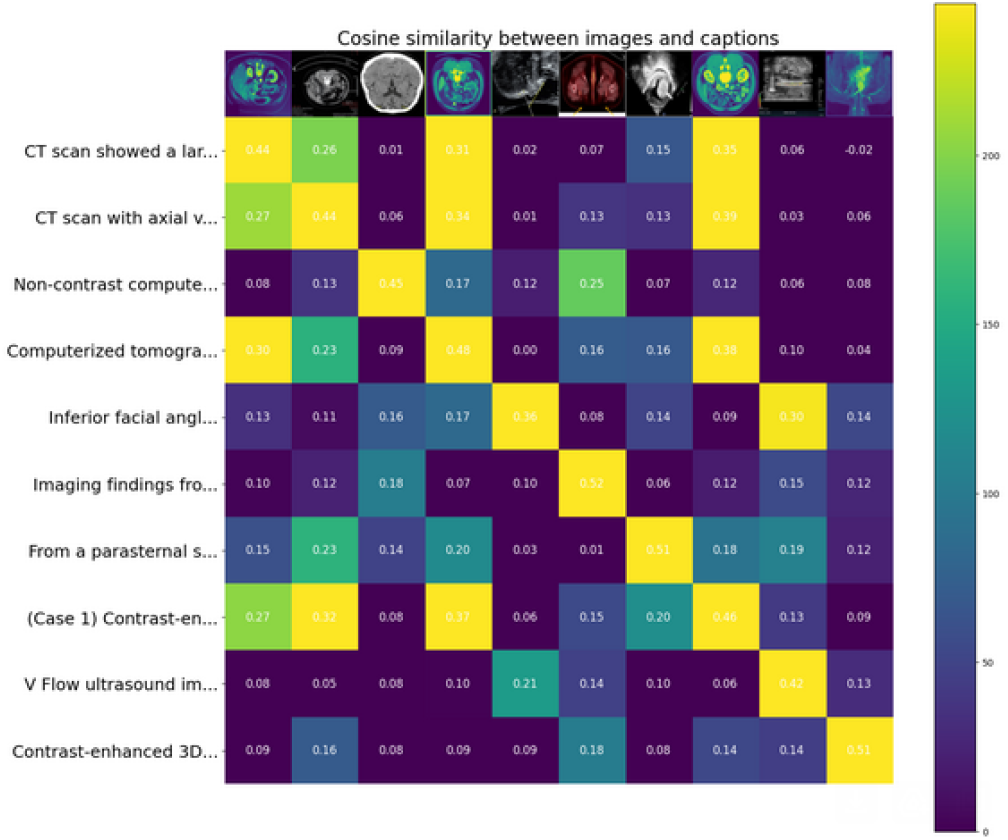
BiomedCLIP-PubMedBERT model Performance Evaluation using Cosine Similarity between Images and Captions from ROCO V2.

#### Algorithm 2

BioMEDCLIPBERT Model Evaluation

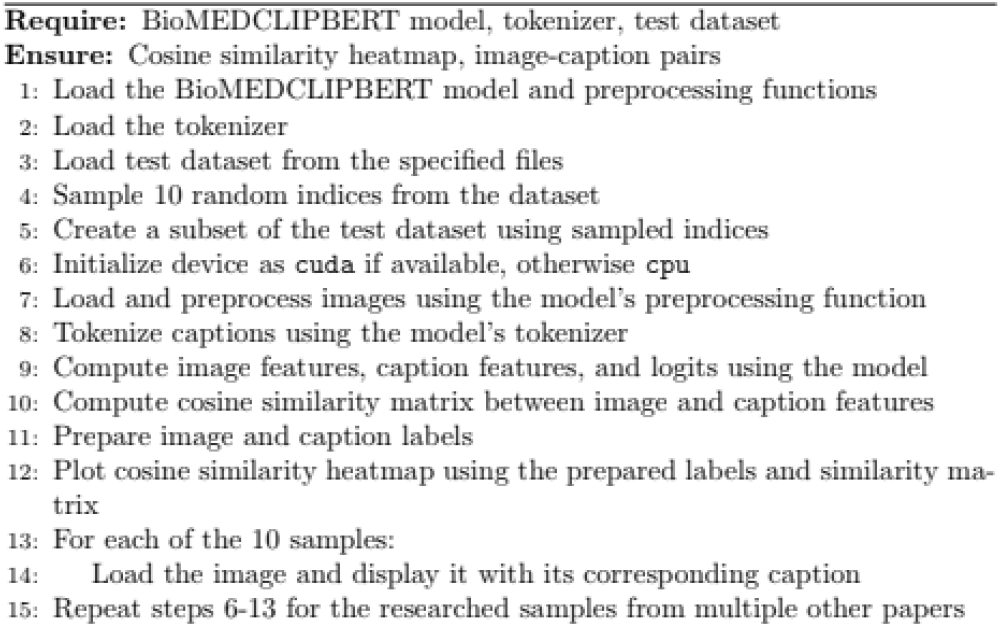

**Table 2:** Randomized Cases of Images and their Captions from ROCO v2 Dataset.

|  | Caption | Image |
| --- | --- | --- |
| 0 | CT scan showed a large stone causing gastric-o... | ROCOv2_2023_test_000357.jpg |
| 1 | CT scan with axial view showing a heterogenous... | ROCOv2_2023_test_009531.jpg |
| 2 | Non-contrast computed tomography of the head w... | ROCOv2_2023_test_008028.jpg |
| 3 | Computerized tomography scan of the abdomen an... | ROCOv2_2023_test_007965.jpg |
| 4 | Inferior facial angle (IFA) measurement. | ROCOv2_2023_test_009484.jpg |
| 5 | Imaging findings from whole body PET CT. On fu... | ROCOv2_2023_test_000558.jpg |
| 6 | From a parasternal short-axis view, the transt... | ROCOv2_2023_test_003880.jpg |
| 7 | (Case 1) Contrast-enhanced computed tomography... | ROCOv2_2023_test_000337.jpg |
| 8 | V Flow ultrasound image of a canine femoral ar... | ROCOv2_2023_test_008248.jpg |
| 9 | Contrast-enhanced 3D MR angiography. Forty-yea... | ROCOv2_2023_test_006850.jpg |

However, since BiomedCLIP-PubMedBERT model is tested in ROCO V2 cases, one may point that this model is a variant of PubMedCLIP [25] which originally trained on ROCO and thus the similarity results may be bias. For this reason we collected random ten images and their captions from ten published articles from PubMed and run the similarity between captions and images using our BiomedCLIP-PubMedBERT model. Table 3 list the ten cases from PubMed and Figure 3 shows the similarity

**Table 3:** Ten Test Cases from PubMed Articles for BiomedCLIP-PubMedBERT.

|  | Caption | Paper/Article | Figure No. | PMID |
| --- | --- | --- | --- | --- |
| 0 | Radiograph of wrist showing ABC of the distal ... | Bone cysts Unicameral and aneurysmal bone cyst | Fig. 6. Section a. | 25579825 |
| 1 | Ear scapha squamous cell carcinoma. Patient ph... | Skin cancer: findings and role of high-resolut... | Fig. 4. | 31069756 |
| 2 | A 49-year-old female COVID-19 patient presenti... | Chest CT manifestations of new coronavirus dis... | Fig. 6. | 32193638 |
| 3 | 63-year-old woman with advanced adenocarcinoma... | Imaging of Precision Therapy for Lung Cancer: ... | Fig. 2a. | 31385753 |
| 4 | Patient 16. Pre- and post-treatment images in ... | Diffusion imaging in obstructive hydrocephalus | Fig. 4. A. | 12812949 |
| 5 | Hip joint MRI of a patient with first-time fem... | Femoral neck fracture after femoral head necro... | Fig. 3. Every section, want to test for multip... | 37907913 |
| 6 | Histology of orthotopic colon tumors. (A) One ... | Magnetic Resonance Imaging and Bioluminescence... | Fig. 5. All sections again | 30988343 |
| 7 | Unoperated "congenitally corrected" transposit... | Imaging congenital heart disease in adults | Fig. 4. a. | 22723533 |
| 8 | A man in his 60s with gastric cancer (well-dif... | Characteristics and Early Diagnosis of Gastric... | Fig. 2. D. | 32321202 |
| 9 | An 8-year-old boy on steroids with cough and n... | Tuberculosis revisited: classic imaging finding... | Fig. 1 | 37217783 |

**Fig. 3:**
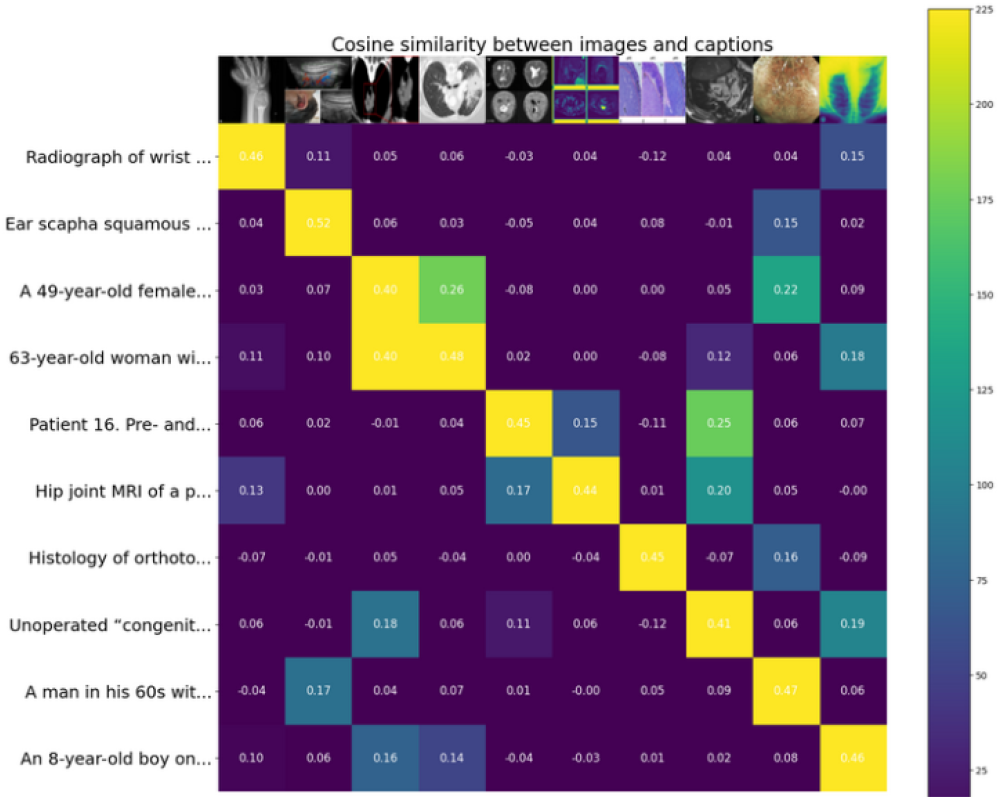
BiomedCLIP-PubMedBERT Performance Evaluation using Cosine Similarity between Images and Captions from Figures Published in PubMed.

## III. Harnessing The full potential of Clinical data Analysis

Understanding and analyzing multimodal clinical data like the electronic healthcare records (EHR) requires not only wider evidence-based support but also the connectivity to the mainstream standards as applied in healthcare systems. Actually analyzing and understanding data requires linkage to the medical concepts repositories like UMLS or SNOMED as well as the support of popular medical vocabulary like MedMentions^6^ corpus which consists of 4,392 papers (Titles and Abstracts) randomly selected from among papers released on PubMed. Additionally the power of understanding and analyzing clinical data requires the power to allow practitioners to interact with the clinical data seamlessly based on sound clinical questioning protocol like PICO^7^ as well as to receive annotations and generate case reports. These added value capabilities for understanding and analyzing clinical data necessitate expanding the machine learning capability of the models used. This means that a machine learning model like our BiomedCLIP-PubMedBERT winning multimodal learning model should prove its applicability beyond the ROCO V2 dataset as well as to integrate the other three non-machine learning capabilities. In this direction we find MedCaT [26] dataset and environment can enable us to achieve this type of integration. MedCat dataset adds on top of 25 K images and captions of ROCO v2, 217,060 figures from 131,410 open access papers as well as 7507 subcaption and subfigure annotations for 2069 compound figures. The wide coverage of evidence-based dataset of MedCat provide another interesting question whether our winning model can span successfully to recognize text and images relationships from MedCat. For this purpose we took another randomized sample from MedCat (see table 4) to test the power of BiomedCLIP-PubMedBERT.

**Table 4:**
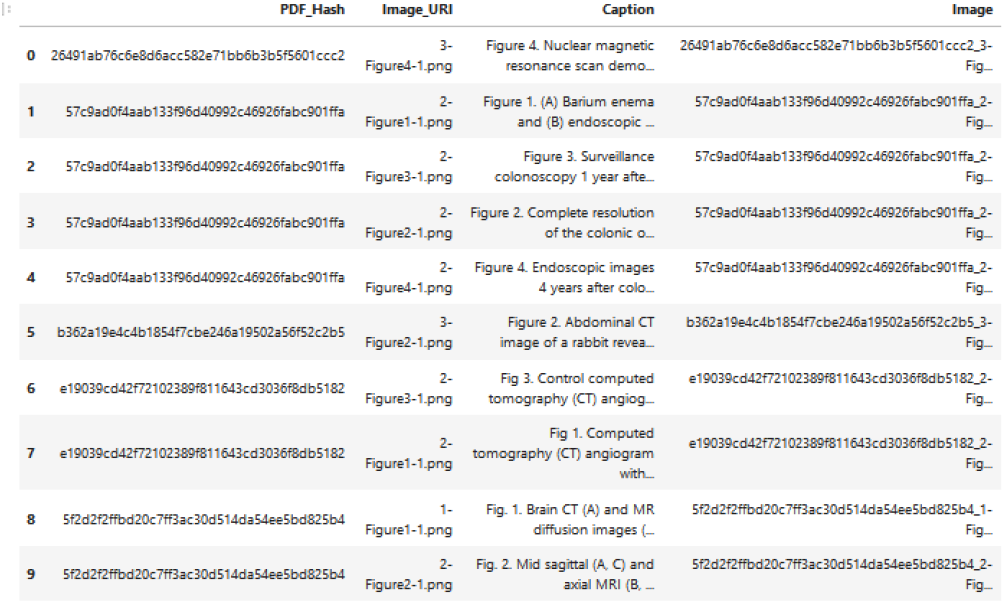
Random Sample from MedCat.

Figure 4 illustrates the cosine similarity of the images and captions using the MedCat random sample. The BiomedCLIP-PubMedBERT model has successfully correlated all the images with their descriptions.

**Fig. 4:**
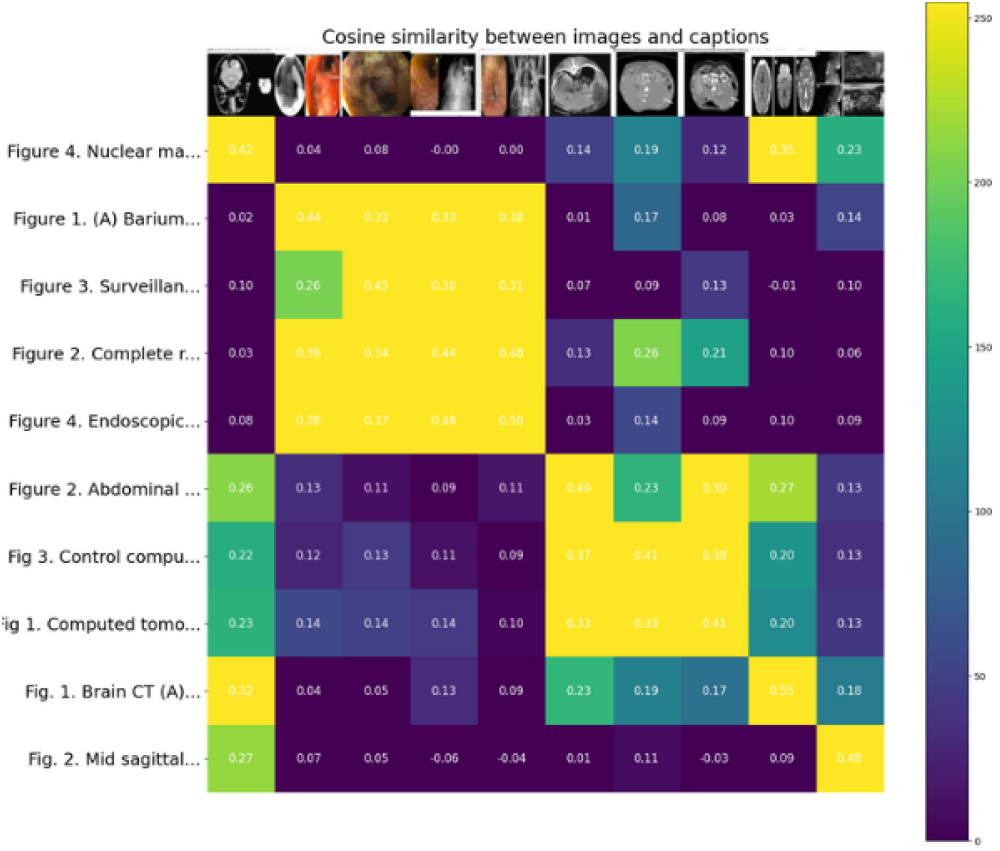
BiomedCLIP-PubMedBERT Performance Evaluation using Cosine Similarity between Images and Captions from MedCat..

## IV. Conclusion

Multimodality has emerged as the primary form of information resource in the paradigm of medical knowledge cognition and reasoning because they can contributes to capturing the underlying complex relationships among clinical and medical elements due to the synergy between the diverse sources [27]. Machine learning based data fusion strategies based on the new wave of meta-transformers are becoming popular approach for modeling these nonlinear relationships echoed from the synergy between the information multimodalities. Although important meta-transformers has been fine-tuned for medical general medical domain, their fine-tuning embeddings can sourced from wide range of publically available data including social media [28]. Hence, these fine-tuned medical meta-transformers cannot be reliably used for evidence-based practice [29]. In our previous research we proved also that evidence-based learning based on normal text analytics transformers that are fine-tuned on evidence-based data cannot provide high accuracy [30]. However, this research article focuses on multimodal learning with Meta-Transformers and their use for clinical support evidence-based medicine. We utilized the TTi-Eval platform to evaluate different fine-tuned medical meta transformers like PubmedCLIP, CLIPMD, BiomedCLIP-PubMedBERT and BioCLIP and found that performance of BiomedCLIP-PubMedBERT outperforms the other models not only in identifying relations between the different medical modalities like clinical narratives assertions and medical imaging but also to support evidences from sound clinical research as published by PubMed. Our overall approached used in this research is illustrated in Figure 4. The dotted parts represent our ongoing research to support more evidence-based mechanisms for clinical support purposes. The current implemented evidence-level focuses on choosing the effective pre-trained medical meta-transformer that provide robust evidence based information. The dotted line is our extension to add a wrapper to enable practitioner’s posses questions around a clinical case using a protocol like PICO and then the learning model help to generate relevant answers based on knowledge from scholarly medical articles. Our initial attempts stated with expanding the use of the MedCat environment that is used for concept annotation [31] and expand it to generate clinical case reports in response to PICO Queries. The full Python implementation details of our first level investigation are given in the Github of this project where the link is available at the acknowledgment of this article.

**Fig. 4:**
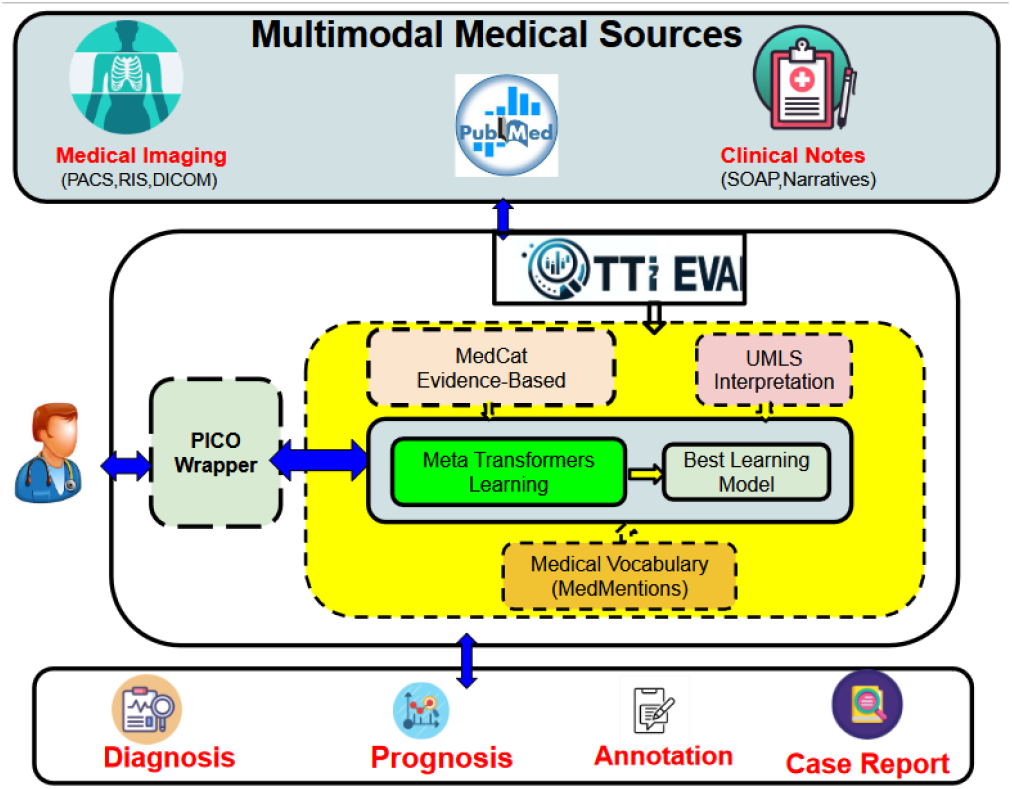
The Overall Platform for Medical Meta-Transformers Learning for Evidence-Based practice.

## Data Availability

All the data and experiments are publicly published in a Github.

https://github.com/TennoSerra/QL4POMR

## Acknowledgment

The first and second authors acknowledge the financial support to this research project from MTACS Accelerates Grant (IT22305-2020) and the first author NSERC DDG Grant (DDG-2021-00014). The experimentation of this project has been published by our MITACS Intern (Abel Serracin Martinez) and it is on the **Github** platform: https://github.com/TennoSerra/QL4POMR

## Footnotes

2 https://huggingface.co/docs/hub/en/index

3 https://openai.com/index/clip/

4 https://www.bracs.icar.cnr.it/

5 https://github.com/google-research-datasets/wit

6 https://github.com/chanzuckerberg/MedMentions?tab=readme-ov-file

7 https://www.nlm.nih.gov/oet/ed/pubmed/pubmed_in_ebp/02-100.html

## References

[1] Zhang, Yiyuan, Kaixiong Gong, Kaipeng Zhang, Hongsheng Li, Yu Qiao, Wanli Ouyang, and Xiangyu Yue. “Meta-transformer: A unified framework for multimodal learning.” arXiv preprint 2307.10802 (2023).

[2] Munikoti, Sai, Ian Stewart, Sameera Horawalavithana, Henry Kvinge, Tegan Emerson, SANDRA E. Thompson, and Karl Pazdernik. “Generalist Multimodal AI: A Review of Architectures, Challenges and Opportunities.” arXiv preprint 2406.05496 (2024).

[3] Wu, Shengqiong, Hao Fei, Leigang Qu, Wei Ji, and Tat-Seng Chua. “Next-gpt: Any-to-any multimodal llm.” arXiv preprint 2309.05519 (2023).

[4] Alec Radford, Jong Wook Kim, Chris Hallacy, Aditya Ramesh, Gabriel Goh, Sandhini Agarwal, Girish Sastry, Amanda Askell, Pamela Mishkin, Jack Clark, et al. Learning transferable visual models from natural language supervision. In International Conference on Machine Learning, pages 8748–8763. PMLR, 2021.

[5] Jean-Baptiste Alayrac, Jeff Donahue, Pauline Luc, Antoine Miech, Iain Barr, Yana Hasson, Karel Lenc, Arthur Mensch, Katie Millican, Malcolm Reynolds, et al. Flamingo: a visual language model for few-shot learning. arXiv preprint 2204.14198, 2022.

[6] Wenhui Wang, Hangbo Bao, Li Dong, and Furu Wei. Vlmo: Unified vision-language pre-training with mixture-of-modality-experts. arXiv preprint 2111.02358, 2021

[7] Peng Wang, An Yang, Rui Men, Junyang Lin, Shuai Bai, Zhikang Li, Jianxin Ma, Chang Zhou, Jingren Zhou, and Hongxia Yang. Unifying architectures, tasks, and modalities through a simple sequence-to-sequence learning framework. arXiv preprint 2202.03052, 2022

[8] Wenhui Wang, Hangbo Bao, Li Dong, Johan Bjorck, Zhiliang Peng, Qiang Liu, Kriti Aggarwal, Owais Khan Mohammed, Saksham Singhal, Subhojit Som, et al. Image as a foreign language: Beit pretraining for all vision and vision-language tasks. arXiv preprint 2208.10442, 2022

[9] Lee, Soyeon, and Minhyeok Lee. “MetaSwin: a unified meta vision transformer model for medical image segmentation.” PeerJ Computer Science 10 (2024): e1762.

[10] Li, Juncheng, Kaihang Pan, Zhiqi Ge, Minghe Gao, Wei Ji, Wenqiao Zhang, Tat-Seng Chua, Siliang Tang, Hanwang Zhang, and Yueting Zhuang. “Fine-tuning multimodal llms to follow zero-shot demonstrative instructions.” In The Twelfth International Conference on Learning Representations. 2023.

[11] Kline, Adrienne, Hanyin Wang, Yikuan Li, Saya Dennis, Meghan Hutch, Zhenxing Xu, Fei Wang, Feixiong Cheng, and Yuan Luo. “Multimodal machine learning in precision health.” arXiv preprint 2204.04777 (2022).

[12] Zhao, Zihao, Yuxiao Liu, Han Wu, Yonghao Li, Sheng Wang, Lin Teng, Disheng Liu et al. “Clip in medical imaging: A comprehensive survey.” arXiv preprint 2312.07353 (2023).

[13] Pan, Xuran, Tianzhu Ye, Dongchen Han, Shiji Song, and Gao Huang. “Contrastive language-image pre-training with knowledge graphs.” Advances in Neural Information Processing Systems 35 (2022): 22895–22910.

[14] Li, Yangguang, Feng Liang, Lichen Zhao, Yufeng Cui, Wanli Ouyang, Jing Shao, Fengwei Yu, and Junjie Yan. “Supervision exists everywhere: A data efficient contrastive language-image pre-training paradigm.” arXiv preprint 2110.05208 (2021).

[15] Xiang, Suncheng, Dahong Qian, Jingsheng Gao, Zirui Zhang, Ting Liu, and Yuzhuo Fu. “Rethinking person re-identification via semantic-based pretraining.” ACM Transactions on Multimedia Computing, Communications and Applications 20, no. 3 (2023): 1–17.

[16] Canese, Kathi, and Sarah Weis. “PubMed: the bibliographic database.” The NCBI handbook 2, no. 1 (2013).

[17] Andrade-Miranda, Gustavo, Vincent Jaouen, Olena Tankyevych, Catherine Cheze Le Rest, Dimitris Visvikis, and Pierre-Henri Conze. “Multi-modal medical Transformers: A meta-analysis for medical image segmentation in oncology.” Computerized Medical Imaging and Graphics 110 (2023): 102308.

[18] Song, Haoyu, Li Dong, Wei-Nan Zhang, Ting Liu, and Furu Wei. “Clip models are few-shot learners: Empirical studies on vqa and visual entailment.” arXiv preprint 2203.07190 (2022).

[19] Alfasly, Saghir, Peyman Nejat, Sobhan Hemati, Jibran Khan, Isaiah Lahr, Areej Alsaafin, Abubakr Shafique et al. “Foundation models for histopathology—fanfare or flair.” Mayo Clinic Proceedings: Digital Health 2, no. 1 (2024): 165–174.

[20] Han, Xu, Zhengyan Zhang, Ning Ding, Yuxian Gu, Xiao Liu, Yuqi Huo, Jiezhong Qiu et al. “Pre-trained models: Past, present and future.” AI Open 2 (2021): 225–250.

[21] Pellegrini, Chantal, Matthias Keicher, Ege Özsoy, Petra Jiraskova, Rickmer Braren, and Nassir Navab. “Xplainer: From x-ray observations to explainable zero-shot diagnosis.” In International Conference on Medical Image Computing and Computer-Assisted Intervention, pp. 420–429. Cham: Springer Nature Switzerland, 2023.

[22] Pellegrini, Chantal, Matthias Keicher, Ege Özsoy, Petra Jiraskova, Rickmer Braren, and Nassir Navab. “Xplainer: From x-ray observations to explainable zero-shot diagnosis.” In International Conference on Medical Image Computing and Computer-Assisted Intervention, pp. 420–429. Cham: Springer Nature Switzerland, 2023.

[23] Javed, Sajid, Arif Mahmood, Iyyakutti Iyappan Ganapathi, Fayaz Ali Dharejo, Naoufel Werghi, and Mohammed Bennamoun. “CPLIP: Zero-Shot Learning for Histopathology with Comprehensive Vision-Language Alignment.” In Proceedings of the IEEE/CVF Conference on Computer Vision and Pattern Recognition, pp. 11450–11459. 2024.

[24] Rückert, Johannes, Louise Bloch, Raphael Brüngel, Ahmad Idrissi-Yaghir, Henning Schäfer, Cynthia S. Schmidt, Sven Koitka et al. “ROCOv2: Radiology Objects in COntext Version 2, an Updated Multimodal Image Dataset.” Scientific Data 11, no. 1 (2024): 688.

[25] Dao, Hong N., Tuyen Nguyen, Cherubin Mugisha, and Incheon Paik. “A Multimodal Transfer Learning Approach using PubMedCLIP for Medical Image Classification.” IEEE Access (2024).

[26] Kraljevic, Zeljko, Thomas Searle, Anthony Shek, Lukasz Roguski, Kawsar Noor, Daniel Bean, Aurelie Mascio et al. “Multi-domain clinical natural language processing with MedCAT: the medical concept annotation toolkit.” Artificial intelligence in medicine 117 (2021): 102083.

[27] Acosta Julián N., Guido J. Falcone, Pranav Rajpurkar, and Eric J. Topol. “Multimodal biomedical AI.” Nature Medicine 28, no. 9 (2022): 1773–1784.

[28] Pourkeyvan, Alireza, Ramin Safa, and Ali Sorourkhah. “Harnessing the power of hugging face transformers for predicting mental health disorders in social networks.” IEEE Access 12 (2024): 28025–28035.

[29] Björklund, Maria, Maria Thereza Perez, Sara Regnér, and Martin Garwicz. “Learning progression from basic scientific scholarship to evidence-based medicine: a multimodal approach.” (2020).

[30] Mohammed, Sabah, Jinan Fiaidhi, and Hashmath Shaik. “Empowering Transformers for Evidence-Based Medicine.” medRxiv (2023): 2023–12.

[31] Kraljevic, Zeljko, Thomas Searle, Anthony Shek, Lukasz Roguski, Kawsar Noor, Daniel Bean, Aurelie Mascio et al. “Multi-domain clinical natural language processing with MedCAT: the medical concept annotation toolkit.” Artificial intelligence in medicine 117 (2021): 102083. https://github.com/CogStack/MedCAT?tab=readme-ov-file

